# Otologic disease among patients with primary ciliary dyskinesia - an international observational study

**DOI:** 10.1101/2022.11.16.22282340

**Authors:** Myrofora Goutaki, Yin Ting Lam, Mihaela Alexandru, Andreas Anagiotos, Miguel Armengot, Mieke Boon, Andrea Burgess, Nathalie Caversaccio, Suzanne Crowley, Sinan Ahmed D. Dheyauldeen, Nagehan Emiralioglu, Ela Erdem, Christine van Gogh, Onder Gunaydın, Eric G. Haarman, Amanda Harris, Isolde Hayn, Hasnaa Ismail-Koch, Bulent Karadag, Céline Kempeneers, Sookyung Kim, Natalie Lorent, Ugur Ozcelik, Charlotte Pioch, Anne-Lise ML Poirrier, Ana Reula, Jobst Roehmel, Panayiotis Yiallouros, Ali Cemal Yumusakhuylu, Jean-François Papon

## Abstract

**Importance:** Otologic disease is common among people with primary ciliary dyskinesia, yet little is known about its spectrum and severity.

**Objective:** We characterized otologic disease among participants with primary ciliary dyskinesia using data from the Ear-Nose-Throat Prospective International Cohort of PCD patients (EPIC-PCD).

**Design:** Cross-sectional analysis of baseline cohort data.

**Setting:** Twelve specialized primary ciliary dyskinesia centers in 10 countries.

**Participants:** We prospectively included children and adults with primary ciliary dyskinesia diagnoses, routine ENT examinations, and completed symptom questionnaires at the same visit or within 2 weeks.

**Exposures:** Potential risk factors associated with increased risk of ear disease.

**Main outcomes and measures:** We describe the prevalence and characteristics of patient-reported otologic symptoms and findings from otologic examinations; we identify potential factors associated with increased risk of ear inflammation and hearing impairment.

**Results:** We included 397 (211 males) participants with median age 15 (range 0–73). A total of 204 (51%) reported ear pain, 110 (28%) ear discharge, and 183 (46%) hearing problems. Adults reported ear pain and hearing problems more frequently when compared with children.

Otitis media with effusion—usually bilateral—from otoscopy was most common among 121 (32%) of 384 participants. Retracted tympanic membrane and tympanic sclerosis were more commonly seen among adults. Tympanometry was performed on 216 participants and showed pathologic type B results for 114 (53%). Audiometry was performed on 273 participants and showed hearing impairment in at least 1 ear, most commonly mild.

Season of visit was the strongest risk factor for problems related to ear inflammation (autumn compared with spring odds ratio, 95% confidence interval: 2.4, 1.5–3.8) and age 30 and older of hearing impairment (age 41–50 compared with age 10 years and younger odds ratio, 95% confidence interval: 3.3, 1.1–9.9).

**Conclusion and relevance:** Many people with primary ciliary dyskinesia suffer from ear problems yet frequency varies, highlighting disease expression differences and possible clinical phenotypes. Understanding differences in otologic disease expression and progression during lifetime may inform clinical decisions about follow-up and medical care. We recommend multidisciplinary primary ciliary dyskinesia management includes regular otologic assessments for all ages even without specific complaints.

**Key Points:** *Question:* What are the characteristics of otologic disease among patients with primary ciliary dyskinesia (PCD)?

*Findings:* Baseline data from a large multicenter cohort of patients with PCD showed frequent reports of ear pain and reduced hearing with age as the main factor associated with hearing impairment. Otitis media with effusion was the most common otoscopic finding; adults often presented with tympanic sclerosis following history of previous ear infections.

*Meaning:* Since otologic disease is an important yet underreported part of PCD’s clinical expression, we recommend otologic assessments for all age groups as part of regular clinical follow-up.

## INTRODUCTION

Primary ciliary dyskinesia (PCD) is a rare inherited disease when pathogenic mutations in disease-causing genes affect ciliary structure or function.^1^ Motile ciliary dysfunction results in a wide range of symptoms from different organ systems.^2–5^ Although the clinical phenotype is heterogeneous, PCD most commonly affects upper and lower airways since ciliary motility is crucial for clearing respiratory secretions.^6–10^ During childhood, many patients with PCD experience recurrent episodes of acute otitis media from defective ciliary function in the Eustachian tube and middle ear, which impair mucociliary clearance and predispose to repeated bacterial infections.^11,12^ Many patients develop bilateral otitis media with effusion (OME) as the disease progresses, yet the prevalence of OME varies between studies.^13,14^ In many otherwise healthy children, OME resolves spontaneously by age 8, but persists beyond this age among children with PCD and needs active management.^15–21^ Indeed, recurrent otitis media and OME results in conductive hearing loss.^16,22^ Developing severe hearing impairment early in life is also complicated by delayed speech-language development.^23^

Retrospective chart review studies of children provide most current knowledge about PCD-related ear problems; however, test result and symptom records are not standardized.^24–26^ In these studies, acute ear problems appear to improve with age—probably because of Eustachian tube anatomical changes and its changing angle with respect to the base of the skull. In an earlier French study, although acute otitis media improved with age, OME reportedly remained frequent among adults and showed no spontaneous improvement.^13^ Even so, patients with specific ultrastructural defects were reported to have a higher prevalence of recurrent otitis media.^13^ However, little is known about age-related changes, such as progression of hearing loss during lifetime or risk factors possibly associated with increased frequency of symptoms.^13,16^

Using data from a large, prospective international cohort, we characterized otologic disease among patients with PCD. Specifically, we describe the prevalence and characteristics of patient-reported otologic symptoms and findings from otologic examinations of children and adults with PCD, and we identify potential factors associated with increased risk of ear disease, specifically ear inflammation and hearing impairment.

## METHODS

### Study design and study population

We analyzed data from the ear-nose-throat (ENT) prospective international cohort of patients with PCD (EPIC-PCD)—an observational multicenter clinical cohort set up in February 2020 hosted at the University of Bern (clinicaltrials.gov identifier: NCT04611516).^27^ EPIC-PCD includes clinical information about patients with PCD of all ages diagnosed by European Respiratory Society (ERS) guidelines and followed at participating centers.^28^ We excluded patients without at least 1 ENT follow-up visit during a year-long period. For this analysis, we included cross-sectional baseline data from all enrolled participants with clinical examinations by ENT specialists and completed symptom questionnaires at the same visit or within 2 weeks with data entered in the study database by July 31, 2022.

Human research ethics committees for all participating centers reviewed and approved EPIC-PCD according to local legislation. We obtained written informed consent or assent in accordance with national data protection laws from participants 14 years or older (with small variations according to local legislation) or from parents or caregivers for younger participants. We report using the Strengthening the Reporting of Observational Studies in Epidemiology (STROBE) recommendations.^29^

### Patient-reported symptoms

Participants or parents completed the standardized, PCD-specific FOLLOW-PCD questionnaire, which is part of the FOLLOW-PCD data collection form, during their scheduled follow-up visit at participating centers.^30^ The FOLLOW-PCD questionnaire includes detailed questions about frequency and characteristics of upper and lower respiratory symptoms during the past 3 months and health-related behaviors, such as active and passive smoking and living environment. There are age-specific questionnaire versions for adults, adolescents 14–17 years, and parents/caregivers of participants 14 years and younger. The FOLLOW-PCD questionnaire was originally developed in English, German, and Greek then translated into the languages of participating centers using a standard procedure.

For otologic symptoms, the questionnaire specifically asks about ear pain, ear discharge, and hearing problems. Questions about symptom frequency are based on a 5-point Likert scale: daily, often, sometimes, rarely, and never. We also asked if each reported symptom was unilateral or bilateral and inquired about ENT symptom seasonal variation. We recorded missing answers as “don’t know” or “never” depending on available answer categories for questions.

### ENT examinations

For planned, clinical reasons, regardless of study participation, participants underwent ENT assessments as part of scheduled follow-up visits at participating centers. ENT specialists assessed ears by otoscopy, tympanometry, and audiometry. We recorded use of hearing aids and presence of tympanostomy tubes. We recorded tympanometry results using Jerger’s description of tympanogram type: type A as normal middle ear status; type B perhaps indicative of OME, tympanic perforation, or sclerosis; types AD and AS as increased and decreased membrane mobility, respectively; and type C as evidence of negative pressure in the middle ear—usually signaling retracted membrane.^31^ We recorded audiometry results using type and World Health Organization (WHO) hearing loss grades.^32^ Since EPIC-PCD is observational, embedded in routine clinical care and management and follows local procedures and protocols, no additional examinations were performed for the purposes of the study. As a result, some assessments were unavailable for participants. Local ENT specialists determined if tympanometry and audiometry were necessary. Using the ENT module from the FOLLOW-PCD form, ENT examinations were recorded in a standardized way.^30^ We recorded and present missing information from ENT assessments as missing data.

### Medical history and other relevant data

We extracted and recorded detailed diagnostic information and information on situs abnormalities and cardiac defects from medical charts at baseline using the corresponding module from FOLLOW-PCD. We entered all data in the study database using Research Electronic Data Capture (REDCap) hosted by the Clinical Trials Unit at the University of Bern.^33^

### Data analysis

We described study population characteristics, prevalence and frequency of patient-reported symptoms, and findings from ENT examinations for the whole cohort and separately for the following age groups: 0–6, 7–14, 15–30, 31–50, and 50 years and older using median and interquartile range for continuous variables and numbers and proportions for categorical variables. We compared prevalence and frequency of symptoms and prevalence of clinical findings between males and females and by age using Chi-squared and t-tests.

We created 2 composite outcome scores representing ear disease: problems related to ear inflammation (*ear inflammation score*) and hearing impairment (*hearing score*). For the ear inflammation score, we included: 1) any reported ear pain or ear discharge, 2) presence of tympanostomy tubes, 3) otitis media, and 4) tympanic perforation during otoscopy; we scored each as either 0 (absence) or 1 (presence) and total score ranged from 0 to 4. For the hearing score, we included: 1) reported hearing problems (0 to 4: never to daily) and 2) audiometry results (0 to 4: normal to profound hearing impairment) with the total score ranging from 0 to 8. We assessed potential factors associated with increased risk of higher ear inflammation or hearing scores using multivariable, ordinal logistic regression models, considering age, age at diagnosis, sex, study center, smoking exposure, season of completed questionnaires, frequency of nasal symptoms reported in questionnaires, and presence of nasal polyps during nasal examinations. We selected factors included in the model based on discussions with clinical specialists and data availability by using directed acyclic graphs. Since age at diagnosis showed strong collinearity with age, we could not include both variables in our main models. We tested if including age at diagnosis instead of age made any difference; since there were no differences, we kept age in our final models. After exploring linear and non-linear effects of age as a continuous variable, we included age groups at ten-year intervals. Due to sample size restrictions, we could not include centers in our final analyses. We therefore ran separate models, including study center as the only explanatory variable. Lastly, in a subgroup of participants with transmission electron microscopy (TEM) results, we repeated both regression models and included only age and ciliary ultrastructural defects to assess if ciliary ultrastructural defect was a risk factor for ear problems. We performed all analyses using Stata (version 15; StataCorp, TX, USA).

## RESULTS

### Demographic, diagnostic and past history characteristics

Participants in our study represent 12 centers from 10 countries. In total, 505 patients were asked to participate in EPIC-PCD of whom 448 (89%) agreed and enrolled in the cohort. From data entered in the database by July 31, 2022, 397 were eligible to participate in this study (eFigure 1). Their median age was 15.2 years, (range 0.2 to 72.4 years), while 218 (55%) were 18 years or older and 186 (47%) were female (Table 1).

**Table 1:**
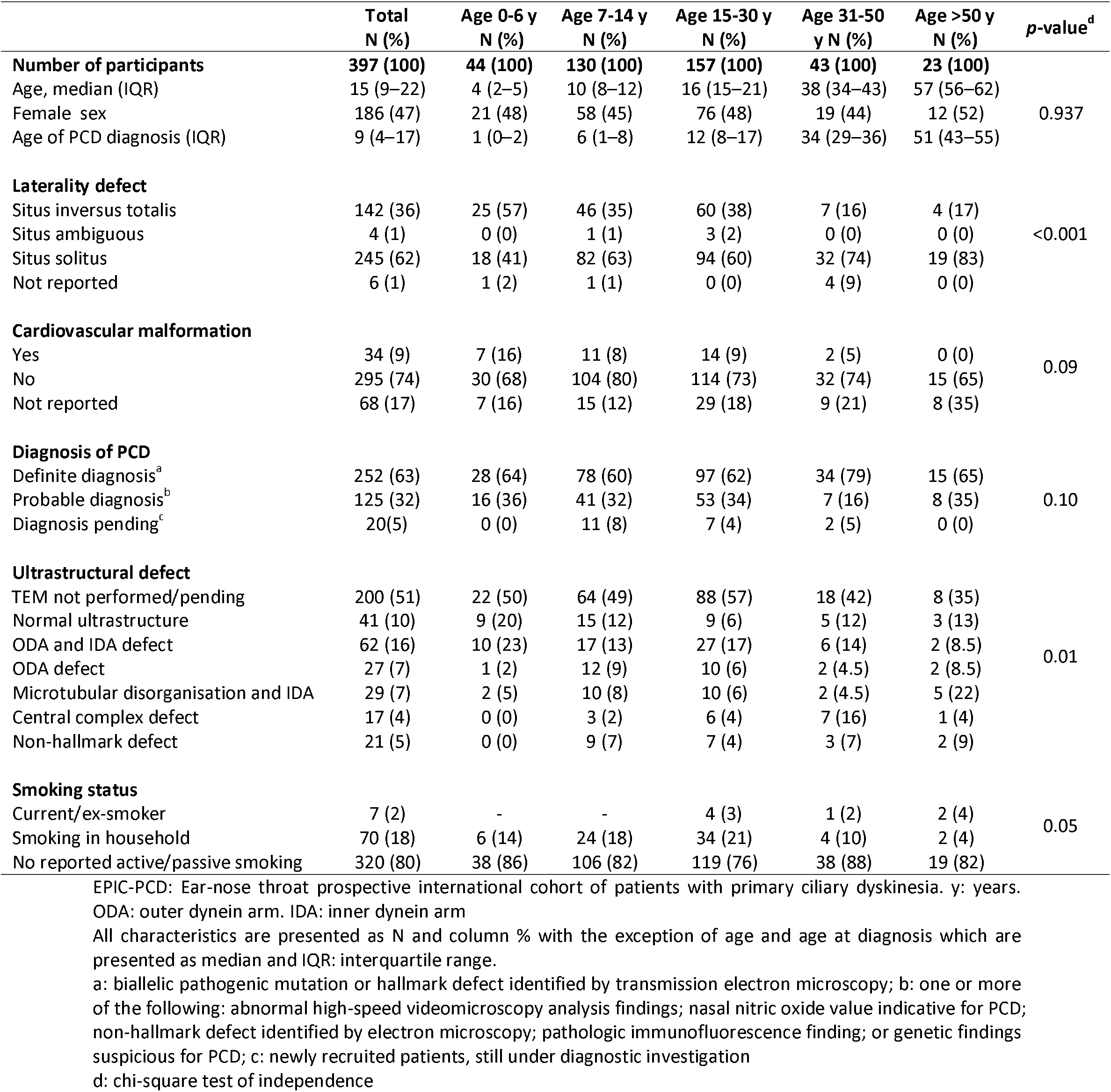
Characteristics of EPIC-PCD participants, overall and by age group (N=397)

In total, 142 (36%) participants had situs inversus and 34 (9%) known cardiac defects. Diagnosis was achieved based on local diagnostic protocols. Nasal nitric oxide (nNO) measurements were performed on 265 (67%) of participants, genetic testing on 281 (71%), TEM on 197 (50%), high-speed videomicroscopy analysis (HSVA) on 227 (57%), and immunofluorescence 72 (18%) (eTables 1-2). Based on ERS guidelines, with biallelic PCD-causing mutation or hallmark defect identified by TEM, definite PCD diagnosis was confirmed for 252 (63%) participants (Table 1).^34^ For remaining participants, 125 (32%) PCD diagnosis was established by combination of several other tests, including HSVA, IF, and nNO, while the remaining 20 (5%) participants were newly diagnosed with strong clinical suspicions and diagnostic results pending at enrolment. Median age at diagnosis was 9 years (range: 0 to 76 years).

### Patient-reported ear symptoms

In total, 204 (51%) participants reported ear pain during the past 3 months, usually bilateral (Figure 1); 52 (13%) participants experienced it daily or often (eTable 3). Unilateral or bilateral ear discharge was reported by 110 (28%) participants; 24 (6%) participants characterized it as daily or often. Hearing problems—mostly bilateral—were reported by 183 (46%) participants. Hearing problems were the most common symptom reported frequently (daily or often) by 75 (19%) participants; 124 (31%) participants reported no ear symptoms (eFigure 2). Most participants reported most troublesome ENT symptoms during winter months, especially during December and January. We found no difference in frequency of reported ear discharge by age (*P*=0.09); however, ear pain (*P*<0.001) and hearing problems (*P*<0.001) were more frequently reported by older participants (eTable 3).

**Figure 1.**
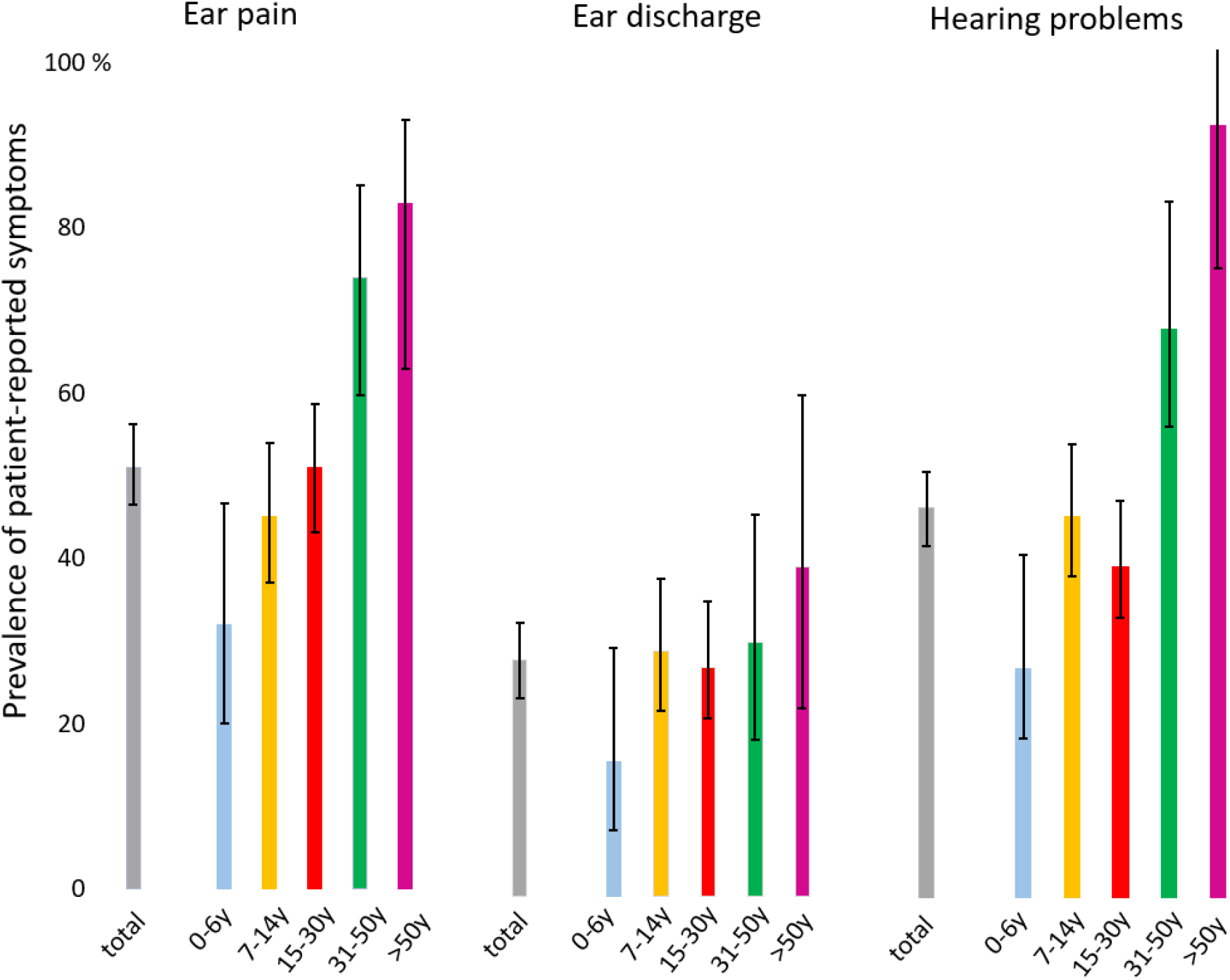
Prevalence of self- and parent-reported ear symptoms of EPIC-PCD participants, overall and by age group (N=387) y: years

### Clinical assessment of the ears

Out of the 397 included participants, 13 (3%) had no otological clinical assessment performed during their follow-up visit at ENT clinics. In Table 2, we present findings from otoscopy for 384 participants. Signs of acute ear disease were rare; 6 (2%) participants had acute otitis media at examination with 9 affected ears. Thirty-eight (10%) participants had active ear discharge from 63 ears and tympanic perforation was recorded for 30 (8%) participants affecting 39 ears. Retracted tympanic membrane was seen in 48 (13%) participants including 75 affected ears. OME—the most common finding—was recorded for 121 (32%) participants with 211 affected ears. Lastly, 69 (18%) participants had signs of tympanic sclerosis with 120 affected ears and 35 (9%) participants had unilateral or bilateral tympanostomy tubes in place at examination (Table 2). From examination findings, retracted membrane and tympanic sclerosis differed by age, and it was more common among adults.

**Table 2:** Otoscopy findings of EPIC-PCD participants, overall and by age group (N=384)

|  | Total<br>N (%) | Age 0-6 y<br>N (%) | Age 7-14 y<br>N (%) | Age 15-30 y<br>N (%) | Age 31-50 y<br>N (%) | Age >50 y<br>N (%) | p-value<sup>a</sup> |
| --- | --- | --- | --- | --- | --- | --- | --- |
| **Number of participants** | 384 (100) | 40 (100) | 126 (100) | 156 (100) | 42 (100) | 20 (100) |  |
| **Acute otitis media** |  |  |  |  |  |  |  |
| Bilateral | 3 (1) | 0 (0) | 3 (2) | 0 (0) | 0 (0) | 0 (0) | 0.73 |
| Unilateral | 3 (1) | 0 (0) | 0 (0) | 3 (2) | 0 (0) | 0 (0) |  |
| No | 364 (95) | 38 (95) | 119 (95) | 146 (94) | 41 (98) | 20 (100) |  |
| Not assessed | 14 (4) | 2 (5) | 4 (3) | 7 (4) | 1 (2) | 0 (0) |  |
| N of affected ears | 9 | 0 | 6 | 3 | 0 | 0 |  |
| **Ear discharge** |  |  |  |  |  |  |  |
| Bilateral | 15 (4) | 1 (2.5) | 7 (5) | 5 (3) | 0 (0) | 2 (10) | 0.75 |
| Unilateral | 23 (6) | 1 (2.5) | 10 (8) | 10 (7) | 1 (2.5) | 1 (5) |  |
| No | 341 (89) | 38 (95) | 107 (85) | 139 (89) | 40 (95) | 17 (85) |  |
| Not assessed | 5 (1) | 0 (0) | 2 (2) | 2 (1) | 1 (2.5) | 0 (0) |  |
| N of affected ears | 63 | 3 | 24 | 20 | 1 | 5 |  |
| **Tympanic perforation** |  |  |  |  |  |  |  |
| Bilateral | 9 (2) | 0 (0) | 3 (2) | 4 (3) | 0 (0) | 2 (10) | 0.39 |
| Unilateral | 21 (6) | 1 (2.5) | 9 (7) | 9 (6) | 2 (5) | 0 (0) |  |
| No | 343 (89) | 36 (90) | 110 (88) | 139 (88) | 40 (95) | 18 (90) |  |
| Not assessed | 11 (3) | 3 (7.5) | 4 (3) | 4 (3) | 0 (0) | 0 (0) |  |
| N of affected ears | 39 | 1 | 15 | 17 | 2 | 4 |  |
| **Retracted membrane** |  |  |  |  |  |  |  |
| Bilateral | 27 (7) | 0 (0) | 7 (5) | 10 (6) | 4 (9.5) | 6 (30) | <0.001 |
| Unilateral | 21 (5) | 2 (5) | 6 (5) | 9 (6) | 1 (2.5) | 3 (15) |  |
| No | 318 (83) | 33 (82.5) | 109 (87) | 129 (83) | 37 (88) | 10 (50) |  |
| Not assessed | 18 (5) | 5 (12.5) | 4 (3) | 8 (5) | 0 (0) | 1 (5) |  |
| N of affected ears | 75 | 2 | 20 | 29 | 9 | 15 |  |
| **Otitis media with effusion** |  |  |  |  |  |  |  |
| Bilateral | 90 (23) | 13 (32.5) | 39 (31) | 26 (17) | 7 (17) | 5 (25) | 0.07 |
| Unilateral | 31 (8) | 2 (5) | 12 (10) | 16 (10) | 0 (0) | 1 (5) |  |
| No | 244 (64) | 22 (55) | 66 (52) | 109 (70) | 34 (81) | 13 (65) |  |
| Not assessed | 19 (5) | 3 (7.5) | 9 (7) | 5 (3) | 1 (2) | 1 (5) |  |
| N of affected ears | 211 | 28 | 90 | 68 | 14 | 11 |  |
| **Tympanic sclerosis** |  |  |  |  |  |  |  |
| Bilateral | 51 (13) | 0 (0) | 9 (7) | 19 (12) | 14 (33) | 9 (45) | <0.001 |
| Unilateral | 18 (5) | 0 (0) | 5 (4) | 12 (8) | 1 (2) | 0 (0) |  |
| No | 277 (72) | 31 (77.5) | 97 (77) | 113 (72) | 25 (60) | 11 (55) |  |
| Not assessed | 38 (10) | 9 (22.5) | 15 (12) | 12 (8) | 2 (5) | 0 (0) |  |
| N of affected ears | 120 | 0 | 23 | 50 | 29 | 18 |  |
| **Tympanostomy tubes** |  |  |  |  |  |  |  |
| Bilateral | 16 (4) | 2 (5) | 6 (5) | 7 (4) | 0 (0) | 1 (5) | 0.40 |
| Unilateral | 19 (5) | 1 (2.5) | 4 (3) | 9 (6) | 4 (10) | 1 (5) |  |
| No | 338 (88) | 34 (85) | 110 (87) | 138 (89) | 38 (90) | 18 (90) |  |
| Not assessed | 11 (3) | 3 (7.5) | 6 (5) | 2 (1) | 0 (0) | 0 (0) |  |
| N of affected ears | 52 | 5 | 16 | 23 | 4 | 3 |  |
| **Hearing aids** | 29 (8) | 2 (5) | 16 (13) | 5 (3) | 2 (5) | 4 (20) | 0.01 |

EPIC-PCD: Ear-nose throat prospective international cohort of patients with primary ciliary dyskinesia. y: years. Findings are presented as N and column %. <sup>a</sup> chi-square test of independence

Tympanometry was performed for 216 (54%) participants. Examinations showed a normal middle ear status (type A) for both ears among 52 (24%) of 216 of participants (Table 3). Type B, which is considered abnormal, was the most common tympanogram for 114 (53%) participants, followed by type C for 34 (16%) participants. Type AD and type AS were seen among 4 (2%) and 33 (15%) participants, respectively.

**Table 3:** Tympanometry findings of EPIC-PCD participants, overall and by age group (N=216)

|  | **Total**<br>N (%) | **Age 0-6 y**<br>N (%) | **Age 7-14 y**<br>N (%) | **Age 15-30 y**<br>N (%) | **Age 31-50 y**<br>N (%) | **Age >50 y**<br>N (%) | **Affected ears**<br>**(N=432)**<br>N (%) |
| --- | --- | --- | --- | --- | --- | --- | --- |
| **Number of participants** | 216 (100) | 14 (100) | 70 (100) | 92 (100) | 26 (100) | 14 (100) |  |
| **Tympanogram type** |  |  |  |  |  |  |  |
| Type A |  |  |  |  |  |  |  |
| Bilateral | 52 (24) | 3 (21) | 14 (20) | 30 (33) | 5 (19) | 0 (0) | 123 (28) |
| Unilateral | 19 (9) | 1 (7) | 6 (9) | 9 (10) | 2 (8) | 0 (0) |  |
| Type B |  |  |  |  |  |  |  |
| Bilateral | 91 (42) | 6 (43) | 38 (54) | 28 (30) | 11 (42) | 8 (57) | 205 (47) |
| Unilateral | 23 (11) | 1 (7) | 5 (7) | 7 (8) | 2 (8) | 4 (29) |  |
| Type AD |  |  |  |  |  |  |  |
| Bilateral | 1 (0) | 0 (0) | 1 (1) | 0 (0) | 0 (0) | 0 (0) | 5 (1) |
| Unilateral | 3 (1) | 1 (7) | 0 (0) | 1 (1) | 1 (4) | 0 (0) |  |
| Type AS |  |  |  |  |  |  |  |
| Bilateral | 19 (8) | 0 (0) | 4 (6) | 13 (14) | 4 (15) | 0 (0) | 52 (12) |
| Unilateral | 14 (6) | 0 (0) | 3 (4) | 6 (7) | 4 (15) | 4 (29) |  |
| Type C |  |  |  |  |  |  |  |
| Bilateral | 19 (9) | 3 (21) | 4 (6) | 8 (9) | 1 (4) | 1 (7) | 53 (12) |
| Unilateral | 15 (7) | 0 (0) | 4 (6) | 3 (3) | 1 (4) | 2 (14) |  |

EPIC-PCD: Ear-nose throat prospective international cohort of patients with primary ciliary dyskinesia. y: years. Tympanogram types by Jerger. Findings are presented as N and column %; categories are not exclusive.

Based on local protocols audiometry testing was performed for 273 (69%) participants and was usually pure tone (75%) or a combination of pure tone, vocal and bone conduction audiometry. In total, based on the WHO hearing loss grading system, 154 (56%) of 273 participants had no impairment (Table 4). Of the remaining participants, most had mild unilateral or bilateral impairment. Five participants had severe and 2 had profound impairment in at least 1 ear. Of all participants, 29 had hearing aids; however, 3 participants refused to wear them (Table 2).

**Table 4:**
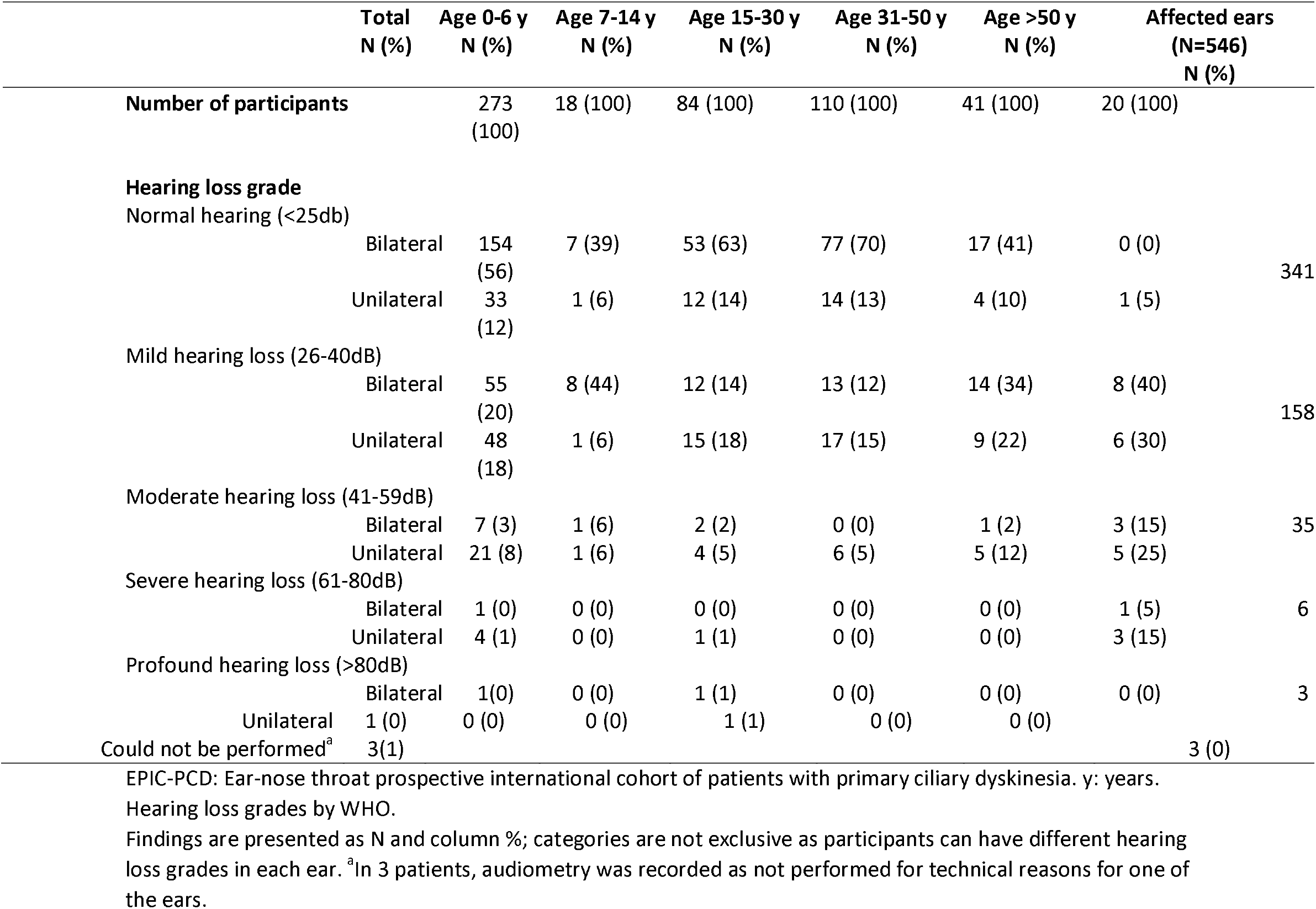
Audiometry findings of EPIC-PCD participants, overall and by age group (N=273)

### Factors associated with ear disease

We found no effect from age, sex, study center, smoking, reported runny or blocked nose, and nasal polyps on the ear inflammation score (Figure 2a). The only factor that showed an association was season of study visit; autumn [odds ratio (OR) and 95% confidence interval (CI): 2.29, 1.44–3.62] showed a higher risk of problems related to ear inflammation compared with spring. For hearing, we found age 30 and older was associated with higher hearing score and the risk increased with age (Figure 2b). Active smoking also showed a strong association (OR 7.9, 95%CI 1.6–39.1 compared with no active or passive smoking); however, the number of smokers participating in the study was very small [7 (2%) participants]. Compared with autumn and winter, participants who visited the clinic in spring or summer had less hearing impairment (Figure 2b). In the subgroup of participants with available TEM results (eFigure 3), we found no associations of specific ultrastructural defects with ear problems.

**Figure 2.**
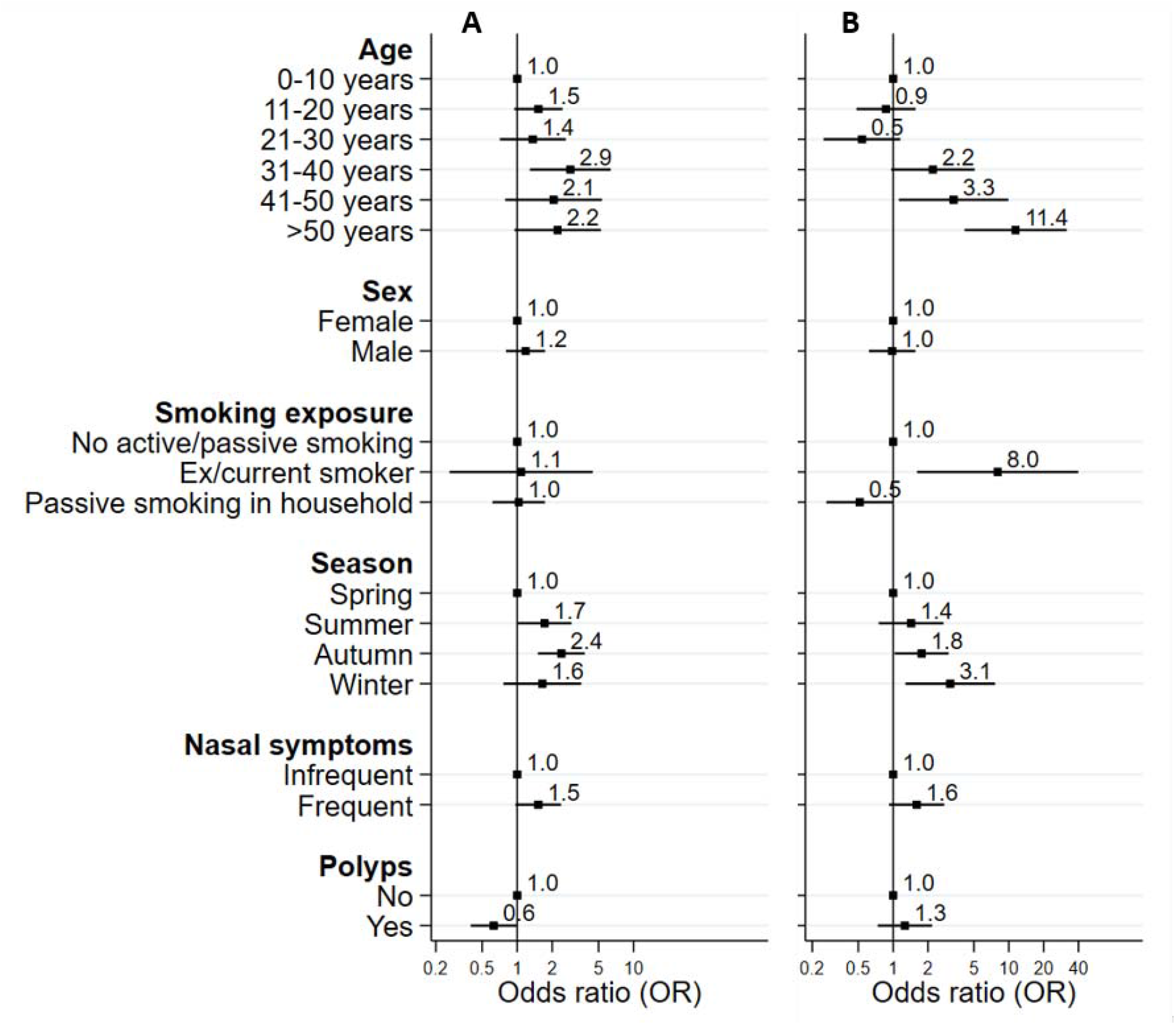
Factors associated with the A) ear infections and B) hearing score among EPIC-PCD participants For the ear infections score, we included any reported ear pain or ear discharge, presence of tympanostomy tubes, otitis media and tympanic perforation during otoscopy, each of which scored as 0 (absence) or 1 (presence); total score ranged from 0 to 4. For the hearing score, we included reported hearing problems (0 to 4: never to daily) and audiometry results (0 to 4: normal to profound hearing impairment) with total score ranging from 0 to 8.

## DISCUSSION

Our study benefitted from standardized acquisition of patient-reported symptoms and clinical assessments to characterize otologic disease in children and adults with PCD. Ear pain and hearing problems were frequently reported, most commonly by adults 30 years and older. OME was the most common finding in ear examination and many adult participants had signs of tympanic sclerosis, a frequent sequelae of recurrent episodes of otitis media, chronic OME, or tympanostomy tubes insertion. Age appeared the main risk factor associated with hearing impairment.

### Strengths and limitations

Our study is the first combining patient-reported symptoms and clinical examination findings of otologic disease among people with PCD; using data from a well-defined, multicenter PCD cohort; and the largest to date focusing on upper airways. Our standardized, PCD-specific questionnaire and ENT evaluation form enhanced collecting high-quality data; it also allows for comparisons with ongoing and future studies using the same tools. Since EPIC-PCD is nested in routine care, we had a high response rate and most invited patients agreed to participate in the study; however, patients with fewer ENT problems might be less willing to participate. Not all assessments were performed for all participants since it was not requested by the study protocol. In turn, it may result in selection bias since tympanometry and audiometry might have been performed most likely for participants with severe ear disease. Since recruitment started in early 2020, our results are possibly affected by the COVID-19 pandemic. Based on anecdotal evidence, patients with PCD suffered from fewer infections due to careful shielding, which possibly led to decreased prevalence of ear problems.^35,36^ As the cohort continues to be followed up longitudinally, it will be important to study possible changes in symptoms and signs coinciding with relaxing pandemic measures. Since we found season a main factor associated with ear inflammation and hearing scores, continued longitudinal follow up also allows us to study seasonal variations of ear disease for longer time periods. Although our study population was large, we still had limited statistical power to study the role of some subgroup characteristics, such as smoking, or different ultrastructural defect and gene groups.

### Comparison with other studies

Since most previous studies were retrospective and information was inconsistently recorded for direct comparisons, prevalence of reported otologic symptoms varied substantially. For instance, in a review summarizing existing literature before 2016, it showed important heterogeneity of study design and population selection, such as reporting a prevalence of hearing impairment from 8% to 100%.^3^ A retrospective study in France included 64 adult patients; it reported hearing loss for 53%, ear pain for 14%, and ear discharge for 8%, while 67% had hearing impairment assessed by audiogram.^37^ Another recent French study showed 71% of 17 adult patients with bronchiectasis and PCD had hearing impairment, 24% conductive.^38^ Older populations and how symptoms were recorded in medical charts possibly explains some differences with our findings. Despite history of recurrent and chronic middle ear disease, there was no record of cholesteatoma for any study participant, which is in accordance with previous literature.^39^ A recent prospective study among 47 children in North America with PCD reported 38% hearing loss and 19% ear pain.^40^ Only including children in the study possibly explains differences with our study, yet a study in the United Kingdom reported abnormal audiometry findings for more than 50% of 271 children.^41^ Prevalence of PCD symptoms may also be underreported by patients and parents because they are accustomed to them. It is also particularly difficult for parents to identify ear symptoms of younger children. It is possible the way patients are asked about specific symptoms during clinical visits may account for some of the recorded differences between studies. In a survey among 74 children and adults with PCD in Switzerland, which also used the FOLLOW-PCD questionnaire, participants reported similar prevalence and frequency of most otologic symptoms compared with our study with just a slightly higher overall prevalence of hearing problems, probably because of older age.^42^ In comparison with 2019 WHO data on hearing impairment that showed 20% of the global population was affected, we found a much higher prevalence of hearing impairment among participants with PCD.^43^

### Conclusion

In addition to respiratory symptoms, many people with PCD suffer from ear problems. Although ear infections are less common in adulthood, sequelae from chronic infections remain and hearing is often impaired, especially among older patients with PCD. Whether a part of the natural disease course or preventable by early, proper management, it remains to be studied. Differences in frequency of ear problems—whether self-reported or identified during clinical examination—highlight differences in disease expression and possibly indicate the existence of clinical phenotypes. Understanding how disease expression differs, how ear, sinonasal, and lower respiratory findings correlate, and how PCD progresses during lifetime, should inform clinical decisions about follow-up and medical care. Since people with PCD underestimate and underreport symptoms, it is possible that many do not receive proper monitoring and management. Therefore, multidisciplinary PCD care needs to include routine otologic and audiologic assessments for patients of all ages even without specific complaints.

## Supporting information

eFigure 1

## Data Availability

All data produced in the present study are available upon reasonable request to the authors

## Author Contributions

M Goutaki developed the concept and designed the study. M Goutaki and YT Lam managed the study and cleaned and standardized the data. M Goutaki performed statistical analyses and drafted the manuscript. All other authors contributed data, interpreted results, and revised critically the manuscript. M Goutaki takes final responsibility for content.

## Funding

The study was supported by a Swiss National Science Foundation Ambizione fellowship (PZ00P3_185923). The authors participate in the BEAT-PCD clinical research collaboration, supported by the European Respiratory Society, and most centers participate in the ERN-LUNG (PCD core).

## Data availability

Data available upon reasonable request.

## Acknowledgments

We want to thank all the people with primary ciliary dyskinesia (PCD) in the cohort and their families, and the PCD support organisations (especially, PCD Family Support Group UK; Association ADCP France; Kartagener Syndrom und Primäre Ciliäre Dyskinesie e. V. Deutschland/ Deutschschweiz; Asociación Nacional de Pacientes con Discinesia Ciliar Primaria DCP España/PCD Spain) for their close collaboration. We also thank all the researchers at participating centers who are involved in enrolment, data collection and data entry, and work closely with us through the whole process of participating in the cohort (all listed below as collaborators of the EPIC-PCD study). We thank Kristin Marie Bivens (ISPM, University of Bern) for her editorial assistance.

P Latzin received grants, honorary, for participation in data safety monitoring board or advisory board from Vertex, Vifor, OM Pharma, Polyphor, Santhera (DMC), Sanofi Aventis within the last 36 months, unrelated to the content of the manuscript. J Roehmel received grants, clinical study recompensations from Vertex, INSMED, Medical Research Council/UK, BMBF, Mukoviszidose Institut within the last 36 months,, unrelated to the content of the manuscript.

## Collaborators of the EPIC-PCD study

(listed in alphabetical order): Dilber Ademhan (Hacettepe University, Turkey), Mihaela Alexandru (AP-HP, France), Andreas Anagiotos (Nicosia General Hospital, Cyprus), Miguel Armengot (La Fe University and Polytechnic Hospital, Spain), Lionel Benchimol (University Hospital of Liège, Belgium), Achim G Beule (University of Münster, Germany), Irma Bon (Vrije Universiteit, the Netherlands), Mieke Boon (University Hospital Leuven, Belgium), Marina Bullo (University of Bern, Switzerland), Andrea Burgess (University of Southampton, UK), Doriane Calmes (University Hospital of Liège, Belgium), Carmen Casaulta (University of Bern, Switzerland), Marco Caversaccio (University of Bern, Switzerland), Nathalie Caversaccio (University of Bern, Switzerland), Bruno Crestani (RESPIRARE, France), Suzanne Crowley (University of Oslo, Norway), Sinan Ahmed. D. Dheyauldeen (University of Oslo, Norway), Sandra Diepenhorst (Vrije Universiteit, The Netherlands), Nagehan Emiralioglu (Hacettepe University, Turkey), Ela Erdem (Marmara University, Turkey), Pinar Ergenekon (Marmara University, Turkey), Nathalie Feyaerts (University Hospital Leuven, Belgium), Gavriel Georgiou (Nicosia General Hospital, Cyprus), Amy Glen (University of Southampton, UK), Christine van Gogh (Vrije Universiteit Amsterdam, the Netherlands), Yasemin Gokdemir (Marmara University, Turkey), Myrofora Goutaki (University of Bern, Switzerland), Onder Gunaydın (Hacettepe University, Turkey), Eric G Haarman (Vrije Universiteit Amsterdam, The Netherlands), Amanda Harris (University of Southampton, UK), Isolde Hayn (Charité-Universitätsmedizin Berlin, Germany), Simone Helms (University of Münster, Germany), Sara-Lynn Hool (University of Bern, Switzerland), Isabelle Honoré (RESPIRARE, France), Hasnaa Ismail Koch (University of Southampton, UK), Bülent Karadag (Marmara University, Turkey), Céline Kempeneers (University Hospital of Liège, Belgium), Synne Kennelly (University of Oslo, Norway), Elisabeth Kieninger (University of Bern, Switzerland), Sookyung Kim (AP-HP, France), Panayiotis Kouis (University of Cyprus, Cyprus), Yin Ting Lam (University of Bern, Switzerland), Philipp Latzin (University of Bern, Switzerland), Marie Legendre (RESPIRARE, France), Natalie Lorent (University Hospital Leuven, Belgium), Jane S Lucas (University of Southampton, UK), Bernard Maitre (RESPIRARE, France), Alison McEvoy (University of Southampton, UK), Rana Mitri-Frangieh (RESPIRARE, France), David Montani (RESPIRARE, France), Loretta Müller (University of Bern, Switzerland), Noelia Muñoz (La Fe University and Polytechnic Hospital, Spain), Heymut Omran (University of Münster, Germany), Ugur Ozcelik (Hacettepe University, Turkey), Beste Ozsezen (Hacettepe University, Turkey), Samantha Packham (University of Southampton, UK), Jean-François Papon (AP-HP, France), Clara Pauly (University Hospital of Liège, Belgium), Charlotte Pioch (Charité-Universitätsmedizin Berlin, Germany), Anne-Lise ML Poirrier (University Hospital of Liège, Belgium), Johanna Raidt (University of Münster, Germany), Ana Reula (La Fe University, Spain), Rico Rinkel (Vrije Universiteit Amsterdam, the Netherlands), Jobst Roehmel (Charité-Universitätsmedizin Berlin, Germany), Andre Schramm (University of Münster, Germany), Simone Tanner (Vrije Universiteit, the Netherlands), Guillaume Thouvenin (RESPIRARE, France), Woolf T Walker (University of Southampton, UK), Hannah Wilkins (University of Southampton, UK), Panayiotis Yiallouros (University of Cyprus, Cyprus), Ali Cemal Yumusakhuylu (Marmara University, Turkey), Niklas Ziegahn (Charité-Universitätsmedizin Berlin, Germany).

## Figure legends

**eFigure 1** Flowchart of patients with PCD who were invited and participated in EPIC-PCD and the study

**eFigure 2** Venn diagram showing overlap of self- and parent-reported symptoms of EPIC-PCD participants (N=387)

**eFigure 3** Association of ciliary ultrastructural defect with the A) ear inflammation and B) hearing score among EPIC-PCD participants

## Notes

### Author Declarations

The study has been reviewed and approved by the local Human Research Ethics Committees at every participating centre, based on local legislation. We list below the names of the ethics committees which approved the study and the approval reference numbers, when applicable. A. The following centres have a pre-existing or new ethical approval, which allows the contribution of pseudonymised data to observational collaborative international studies (covers the EPIC-PCD study): 1.University Childrens Hospital Charité-Universitätsmedizin, Berlin, Germany: Ethical Committee Charité (EA2/003/21) 2. University Childrens hospital, Bern, Switzerland: Cantonal Ethics Committee of Bern (KEKBE: 060/2015) 3. University of Cyprus: Ethical Committee for biomedical research in Leukosia Cyprus (EEBK/EΠ/2013/21) 4. Marmara University Istanbul, Turkey: Ethical Committee of Marmara University (09.2018.395) 5. University Hospital of Southampton, United Kingdom: Southampton and South West Hampshire research ethics committee (06/Q1702/109) B. The following centres applied for ethical approval to participate specifically to the EPIC-PCD study: 1. VU University medical center (VUmc), Amsterdam, The Netherlands: The Medical Ethics Review Committee of VU University Medical Center reviewed the application and concluded on 24th of November 2020 that no approval is needed to participate to the EPIC-PCD cohort as the Medical Research Involving Human Subjects Act does not apply to the study. 2. Hacettepe University, Ankara, Turkey: Non-interventional clinical research EC of Hacettepe University (2020/11-47) 3. University Hospital of Leuven, Belgium: Ethical Committee for Research of University Hospitals Leuven (S64411) 4. Hospital Universitario La Fe in Valencia, Spain: Ethical Committee of medical investigations of Hospital Universitario La Fe (2020-498-1) 5. University Hospital Bicetre Paris-Sud, Paris, France: The AP-HP Direction de la Recherche Clinique et de l'Innovation reviewed the application and concluded on 4th of February 2021 that that no approval is needed to participate to the EPIC-PCD cohort as the Jardé law that regulates clinical research in France does not apply to the study.

