## Supplementary material for "Otologic disease among patients with primary ciliary dyskinesia - an international observational study": eFigure 1

This online-only document includes the following elements, in the order that they are cited in the manuscript:

**eFigure 1** Flowchart of patients with PCD who were invited and participated in EPIC-PCD and the study

**eTable 1** Centre and diagnostic information of EPIC-PCD participants, overall and by age group (N=397)

**eTable 2**: Genetic mutations reported in EPIC-PCD participants with biallelic pathogenic variants or compound heterozygosity

**eTable 3**: Frequency of self- and parent-reported ear symptoms of EPIC-PCD participants, overall and by age group (N=387)

**eFigure 2** Venn diagram showing overlap of self- and parent-reported symptoms of EPIC-PCD participants (N=387)

**eFigure 3** Association of ciliary ultrastructural defect with the A) ear infections and B) hearing score among EPIC-PCD participants

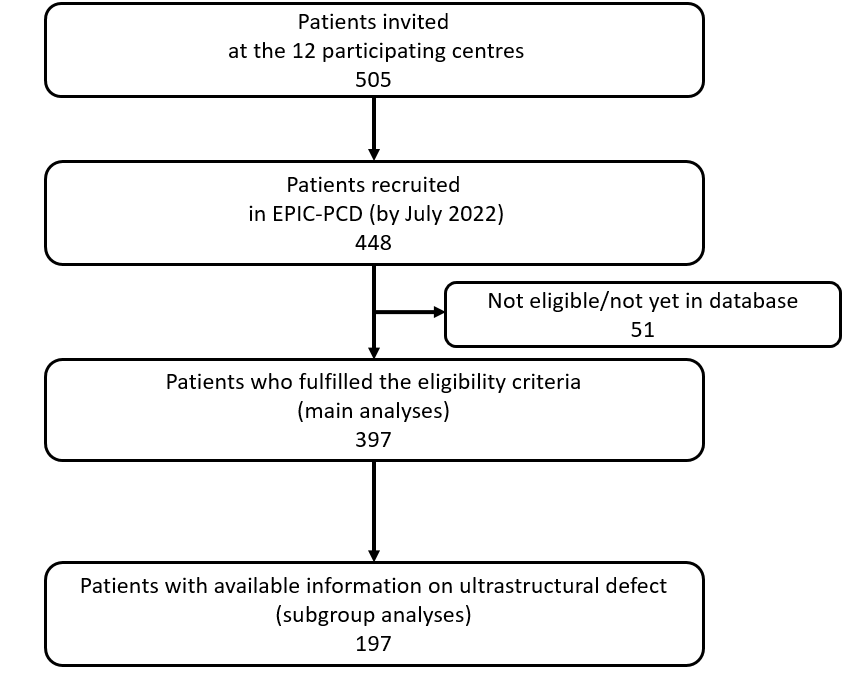

**eFigure 1** Flowchart of patients with PCD who were invited and participated in EPIC-PCD and the study

**eTable 1** Centre and diagnostic information of EPIC-PCD participants, overall and by age group (N=397)

|  | **Total**  **N (%)** | **Age 0-6 y**  **N (%)** | **Age 7-14 y**  **N (%)** | **Age 15-30 y**  **N (%)** | **Age 31-50y N (%)** | **Age >50 y N (%)** |
| --- | --- | --- | --- | --- | --- | --- |
| **Number of participants** | **397 (100)** | **44 (100)** | **130 (100)** | **157 (100)** | **43 (100)** | **23 (100)** |
| **Centre** |  |  |  |  |  |  |
| Amsterdam | 26 (7) | 3 (7) | 12 (9) | 11 (7) | 0 (0) | 0 (0) |
| Ankara | 60 (15) | 5 (11) | 23 (18) | 32 (20) | 0 (0) | 0 (0) |
| Berlin | 43 (11) | 8 (18) | 4 (3) | 16 (10) | 10 (23) | 5 (22) |
| Bern | 7 (2) | 1 (2) | 2 (2) | 4(3) | 0 (0) | 0 (0) |
| Cyprus | 22 (6) | 3 (7) | 3 (2) | 6 (4) | 6 (14) | 4 (17) |
| Istanbul | 66 (17) | 4 (9) | 26 (20) | 34 (22) | 2 (5) | 0 (0) |
| Leuven | 12 (3) | 0 (0) | 5 (4) | 6 (4) | 1 (2) | 0 (0) |
| Liege | 10 (2) | 1 (2) | 4 (3) | 3 (2) | 2 (5) | 0 (0) |
| Oslo | 39 (9) | 3 (7) | 22 (17) | 14 (9) | 0 (0) | 0 (0) |
| Paris | 53 (13) | 0 (0) | 7 (5) | 17 (11) | 20 (47) | 9 (39) |
| Southampton | 43 (11) | 14 (32) | 22 (17) | 7 (4) | 0 (0) | 0 (0) |
| Valencia | 16 (4) | 2 (5) | 0 (0) | 7 (4) | 2 (5) | 5 (22) |
| **Nasal nitric oxide measurement** |  |  |  |  |  |  |
| Indicative for PCD | 249 (63) | 22 (50) | 76 (59) | 103 (65) | 32 (74) | 16 (70) |
| Not indicative for PCD | 16 (4) | 1 (2) | 3 (2) | 9 (6) | 3 (7) | 0 (0) |
| Not performed/pending | 132 (33) | 21 (48) | 51 (39) | 45 (29) | 8 (19) | 7 (30) |
| **Transmission Electron Microscopy** |  |  |  |  |  |  |
| Hallmark defect | 135 (34) | 13 (30) | 42 (32) | 53 (34) | 17 (40) | 10 (43) |
| Other defect | 21 (5) | 0 (0) | 9 (7) | 7 (4) | 3 (7) | 2 (9) |
| Normal ultrastructure | 41 (10) | 9 (20) | 15 (12) | 9 (6) | 5 (11) | 3 (13) |
| Not performed/pending | 200 (51) | 22 (50) | 64 (49) | 88 (56) | 18 (42) | 8 (35) |
| **High-speed videomicroscopy analysis** |  |  |  |  |  |  |
| Indicative for PCD | 207 (52) | 30 (68) | 62 (48) | 79 (51) | 19 (44) | 17 (74) |
| Other/unclear | 13 (3) | 4 (9) | 6 (5) | 2 (1) | 1 (2) | 0 (0) |
| Normal motility | 6 (2) | 1 (2) | 2 (2) | 2 (1) | 1 (2) | 0 (0) |
| Not performed/pending | 170 (43) | 9 (21) | 59 (45) | 74 (47) | 22 (52) | 6 (26) |
| **Genetic testing** |  |  |  |  |  |  |
| No pathogenic variants | 33 (8) | 6 (14) | 12 (9) | 11 (7) | 2 (5) | 2 (9) |
| Results pending | 26 (6) | 1 (2) | 9 (7) | 16 (10) | 0 (0) | 0 (0) |
| Biallellic pathogenic variants/  compound heterozygosity | 181 (46) | 20 (45) | 57 (44) | 65 (42) | 27 (63) | 12 (52) |
| Heterozygous variant | 39 (10) | 3 (7) | 7 (5) | 21 (13) | 4 (9) | 4 (17) |
| Not performed | 118 (30) | 14 (32) | 45 (35) | 44 (28) | 10 (23) | 5 (22) |
| **Immunofluorescence** |  |  |  |  |  |  |
| Indicative for PCD | 36 (9) | 7 (16) | 6 (5) | 21 (13) | 2 (5) | 0 (0) |
| Not indicative for PCD | 36 (9) | 8 (18) | 17 (13) | 9 (6) | 1 (2) | 1 (4) |
| Not performed/pending | 325 (82) | 29 (66) | 107 (82) | 127 (81) | 40 (93) | 22 (96) |

EPIC-PCD: Ear-nose throat prospective international cohort of patients with primary ciliary dyskinesia. y: years.

Characteristics are presented as N and column %.

**eTable 2**: Genetic mutations reported in EPIC-PCD participants with biallelic pathogenic variants or compound heterozygosity

| **Genetic mutation** | **N (%)** |
| --- | --- |
| DNAH5 | 32 (18) |
| DNAH11 | 24 (13) |
| CCDC40 | 21 (11) |
| HYDIN | 13 (7) |
| CCDC39 | 9 (5) |
| RSPH4A | 9 (5) |
| DNAI1 | 9 (5) |
| CCNO | 8 (4) |
| RSPH9 | 8 (4) |
| DNAAF1 | 6 (3) |
| RSPH3 | 5 (3) |
| CCDC114 | 5 (3) |
| DNAH9 | 3 (2) |
| C11ORF70 | 4 (2) |
| TTC25 | 3 (1) |
| DNAI2 | 3 1) |
| DNAAF11 | 3 (1) |
| CCDC65 | 2 (1) |
| DNAFF2 | 2 (1) |
| CCDC103 | 1 (1) |
| ZMYND10 | 1 (1) |
| DRC1 | 1 (1) |
| DNAH1 | 1 (1) |
| DNAAF3 | 1 (1) |
| DNAAF5 | 1 (1) |
| ARMC4 | 1 (1) |
| CCDCC151 | 1 (1) |
| DCLRE1C | 1 (1) |
| Mutation not reported | 3 (1) |
| **Total** | **181 (100)** |

EPIC-PCD: Ear-nose throat prospective international cohort of patients with primary ciliary dyskinesia.

**eTable 3**: Frequency of self- and parent-reported ear symptoms of EPIC-PCD participants, overall and by age group (N=387)

|  | | **Total**  **N (%)** | **Age 0-6 y**  **N (%)** | **Age 7-14 y**  **N (%)** | **Age 15-30 y**  **N (%)** | **Age 31-50 y N (%)** | **Age >50 y N (%)** | ***p*-value^a^** |
| --- | --- | --- | --- | --- | --- | --- | --- | --- |
| **Number of participants** | 397 (100) | | 44 (100) | 130 (100) | 157 (100) | 43 (100) | 23 (100) |  |
| **Ear pain** | |  |  |  |  |  |  | <0.001 |
| Daily | | 14 (3) | 0 (0) | 2 (1) | 6 (4) | 4 (9) | 2 (9) |  |
| Often | | 36 (9) | 2 (4) | 10 (8) | 16 (10) | 3 (7) | 5 (22) |  |
| Sometimes | | 70 (18) | 6 (14) | 25 (19) | 21 (13) | 9 (21) | 9 (39) |  |
| Rarely | | 84 (21) | 6 (14) | 22 (17) | 37 (24) | 16 (37) | 3 (13) |  |
| Never/not reported | | 193 (49) | 30 (68) | 71 (55) | 77 (49) | 11 (26) | 4 (17) |  |
| **Ear discharge** | |  |  |  |  |  |  | 0.09 |
| Daily | | 7 (2) | 1 (2) | 3 (2) | 1 (1) | 1 (2) | 1 (4) |  |
| Often | | 15 (4) | 0 (0) | 2 (2) | 7 (4) | 3 (7) | 3 (13) |  |
| Sometimes | | 40 (10) | 2 (5) | 21 (16) | 13 (8) | 3 (7) | 1 (4) |  |
| Rarely | | 48 (12) | 4 (9) | 12 (9) | 22 (14) | 6 (14) | 4 (17) |  |
| Never/not reported | | 287 (72) | 37 (84) | 92 (71) | 114 (73) | 30 (70) | 14 (61) |  |
| **Hearing problems** | |  |  |  |  |  |  | <0.001 |
| Daily | | 37 (9) | 2 (5) | 10 (8) | 9 (6) | 7 (16) | 9 (39) |  |
| Often | | 38 (10) | 4 (9) | 10 (8) | 10 (6) | 8 (18) | 6 (26) |  |
| Sometimes | | 61 (15) | 5 (11) | 25 (19) | 17 (11) | 9 (21) | 5 (22) |  |
| Rarely | | 47 (12) | 1 (2) | 14 (10) | 26 (16) | 5 (12) | 1 (4) |  |
| Never/not reported | | 214 (54) | 32 (73) | 71 (55) | 95 (61) | 14 (33) | 2 (9) |  |

EPIC-PCD: Ear-nose throat prospective international cohort of patients with primary ciliary dyskinesia. y: years.

Symptoms are presented as N and column %. ^a^ chi-square test of independence

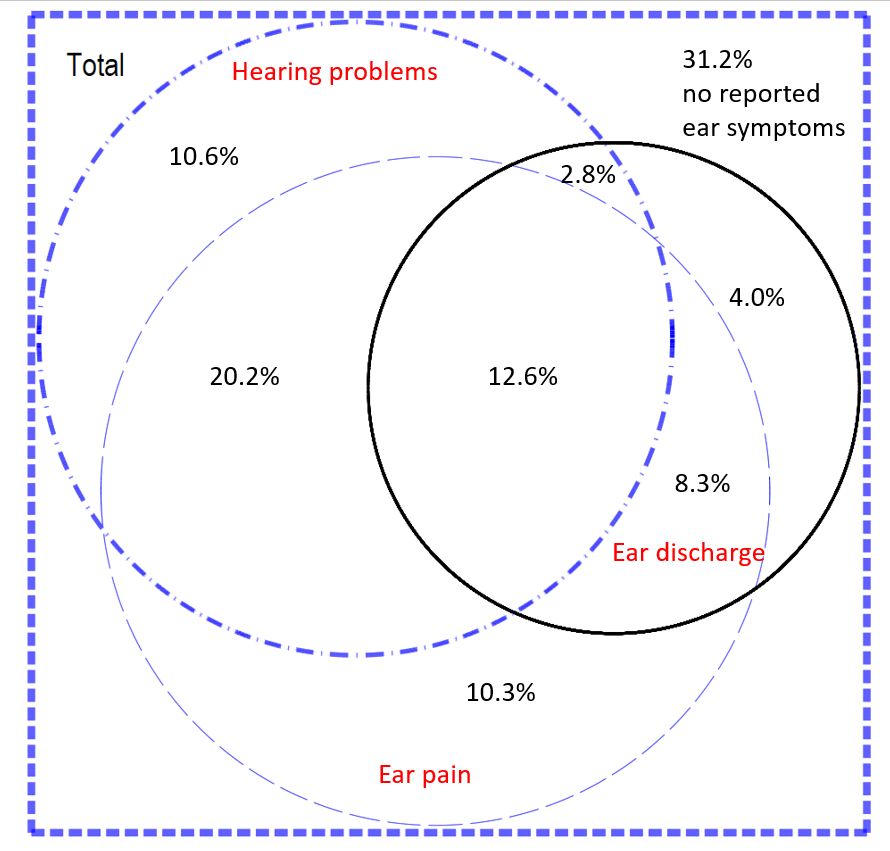
**eFigure 2** Venn diagram showing overlap of self- and parent-reported symptoms of EPIC-PCD participants (N=387)

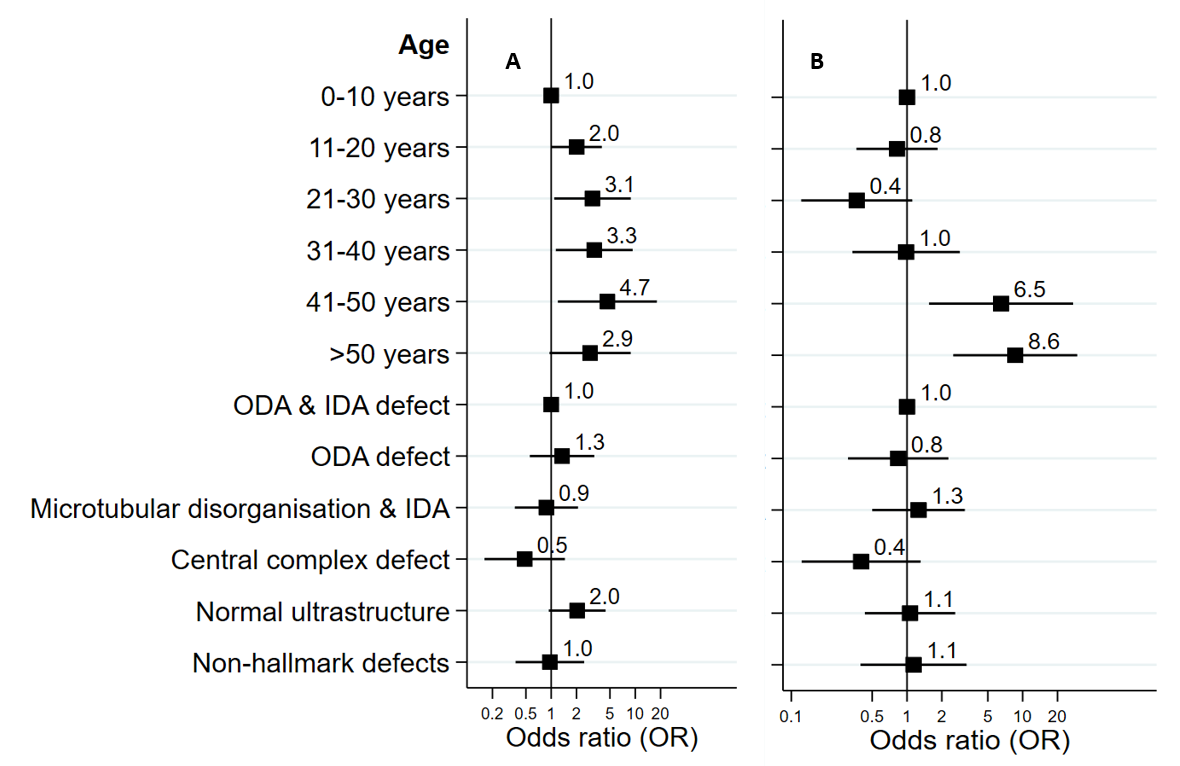

**eFigure 3** Association of ciliary ultrastructural defect with the A) ear infections and B) hearing score among EPIC-PCD participants

For the ear infections score, we included any reported ear pain or ear discharge, presence of tympanostomy tubes, otitis media and tympanic perforation during otoscopy, each of which scored as 0 (absence) or 1 (presence); total score ranged from 0 to 4. For the hearing score, we included reported hearing problems (0 to 4: never to daily) and audiometry results (0 to 4: normal to profound hearing impairment) with total score ranging from 0 to 8.
